# COVID-19 pandemic and changes in urban-rural inequalities of suicide in the Province of Buenos Aires-Argentina, 2017-2021

**DOI:** 10.1101/2025.01.09.25320266

**Authors:** Carlos M. Leveau, Guillermo A. Velázquez

**Affiliations:** Centro de Salud Mental Comunitaria “Mauricio Goldenberg”, Universidad Nacional de Lanús, Argentina; Consejo Nacional de Investigaciones Científicas y Técnicas (CONICET), Argentina; Instituto de Geografía, Historia y Ciencias Sociales, Consejo Nacional de Investigaciones Científicas y Técnicas – Universidad Nacional del Centro de la Provincia de Buenos Aires, Tandil, Argentina

**Keywords:** cities, rural areas, wounds and injuries, violence, mortality

## Abstract

**Introduction:** Our objective was to analyze the urban-rural inequalities of suicide between the pre-pandemic and pandemic periods in the Province of Buenos Aires (PBA), during 2017-2021.

**Methods:** Data regarding sex, age and municipality of occurrence of suicides were used, provided by the Ministry of Health (MSAL) of the PBA and the National Criminal Information System (SNIC). Municipalities were divided into four urbanization categories according to their population density. Annual variations in suicide were analyzed and through two periods: pre-pandemic (2017-2019) and pandemic (2020-2021). Negative binomial regressions were calculated to estimate suicide inequalities between urbanization categories.

**Results:** Men, people 60+ years old, residents of municipalities with high social fragmentation, and with low levels of poverty presented a higher risk of suicide. The year 2020 recorded the lowest risk of suicide. The most urbanized municipalities had a lower risk of suicide compared to the most rural municipalities. With data from the MSAL, these inequalities were similar comparing between the pre-pandemic and pandemic periods. The SNIC data showed some differences possibly attributable to under-reporting of suicides in 2020.

**Conclusion:** Analysis of both data sources suggest that the social and economic effects of the COVID-19 pandemic did not lead to an increase in urban-rural suicide gaps.

## Introduction

Worldwide, suicide constitutes a public health problem, being one of the main causes of premature death and representing 1 in every 100 deaths, according to 2019 estimates ^1^. While the suicide rate has decreased between 2000 and 2019 worldwide, in the region of the Americas this rate has increased. In Argentina, although the suicide rate has shown a downward trend since 2001 ^2^, in the South American context it has the highest rate after Uruguay ^1^.

There is abundant literature, focused on developed countries and condensed in literature reviews ^3,4^, showing a higher risk of suicide in rural areas. In Latin American countries there also seems to be a higher risk of suicide outside large cities, as indicated by studies carried out in Argentina ^5–7^, Colombia ^8^, and Cuba ^9^, although another study in Colombia showed a higher suicide rate in urban areas ^10^. Three types of interrelated factors may be inherent to rural spaces, leading to a greater risk of suicide with respect to large cities: contextual (low access to basic services, concentration of land, drought and floods, exposure to agrochemicals ^11^), collective (values, norms and traditions typical of rural areas, such as the use of firearms), and compositional (demographic structure, types of employment, occupational category) ^3^. With the emergence of the SARS-CoV-2 virus and its rapid spread on a global scale, the first most studied question has been the impact of the COVID-19 pandemic on the temporal variation of suicides. Two competing hypotheses emerged from this question.

First, it was hypothesized that physical-social distancing measures would deteriorate mental health by increasing social isolation, decreasing access to health services, increasing exposure to domestic violence, and generating higher levels of economic uncertainty ^12^. The combination of these factors would lead to an increase in suicides. Second, following Durkheim, social shocks, such as that experienced with the COVID-19 pandemic, could cause greater social integration, reducing the risk of suicide in the population ^13^. However, the evidence so far does not show significant variations in suicide associated with the COVID-19 pandemic ^14^.

When considering an urban-rural geographic gradient, a question arises whether any of these possible explanations could have had greater weight in rural or urban areas. Three factors have been raised that could be exacerbated in rural areas with the emergence of the COVID-19 pandemic ^15^. First, it has been argued that social distancing measures would further increase the levels of social isolation of rural populations, coupled with an increase in levels of domestic violence. Second, greater access to firearms in rural populations would increase the risk of suicide in a context of greater uncertainty and social isolation. Third, a worsening of mental health with the onset of the pandemic could have a more negative impact on rural populations, with less access to mental health services.

Although a greater vulnerability to suicide has been suggested in rural populations with the onset of the COVID-19 pandemic, the few published studies do not provide conclusive evidence in favor of this hypothesis. In the United States, an analysis of suicide between 2019 and 2020 showed significant declines in rates in larger metropolitan areas, while rates in non-metropolitan counties remained stable ^16^. In Mexico, less densely populated states tended to record fewer suicides than expected during 2020 ^17^. Similarly, in Ecuador the proportion of suicides in rural areas was significantly lower, compared to urban areas, during the pandemic period (March 2020 to June 2021) compared to the pre-pandemic period (January 2015 to February 2020) ^18^. In this study we take the Province of Buenos Aires (PBA) as the study area, the most populated jurisdiction in Argentina and with great geo-demographic contrasts, between its largely rural western part and the mega-city Buenos Aires in the northeast. Therefore, the objective of this work is to analyze the urban-rural inequalities of suicide between the pre-pandemic and pandemic periods in the PBA, during 2017-2021.

## Methods

This is a retrospective ecological study, consisting of multivariate quantitative analysis of secondary data. Due to the latter, approval of the present study by an Ethics Committee was not necessary. Data on deaths by suicide were used during the five-year period 2017-2021, from two data sources. First, we used mortality data provided by the Ministry of Health of the province of Buenos Aires (hereinafter, MSAL) ^19^, from which we analyzed deaths coded as X60-84 according to the ICD-10. Secondly, data from the National Criminal Information System (SNIC) was used, consolidated and processed by the National Directorate of Criminal Statistics (DNEC), dependent on the Ministry of National Security ^20^. The SNIC compiles data collected by provincial and national security forces throughout the entire Argentine territory. It is the members of these forces who determine the type of death (road accident, homicide, or suicide) according to the information they collect at the scene. In both data sources, deaths were disaggregated by age, sex, and municipality of suicide occurrence. Age was considered as a categorical variable (10-29, 30-59, and 60 or older) in the regression models. The study population included the 134 municipalities that make up the PBA. Municipalities are political-administrative territorial subdivisions, also called parties. According to provisional data from the 2022 Population Census, the PBA represents 38% (17.6 million) of the Argentine population. The annual population of each municipality during 2017-2021, disaggregated by age and sex, was estimated through linear projections using data from the 2001 and 2010 population censuses because said data corresponding to the 2022 Census have not yet been published.

Due to overdispersion of data ^21^, negative binomial regression models were used to test the relationship between different levels of urbanization and suicide risk during 2017-2021. Sex, age group, and year of suicide occurrence were used as independent variables, while the municipalities of the PBA were divided into urbanization quartiles according to their population density (inhabitants/kilometer2). According to Tisdale ^22^, urbanization is population concentration, so the most urbanized municipalities are those with the highest population density, while the most rural municipalities are the ones with the lowest population density. In addition, poverty and social fragmentation indices were calculated for each municipality. Population density and poverty and social fragmentation indices were calculated using data from the 2010 Census. The poverty index, modified from that calculated by Carstairs ^23^, was the result of the sum of the z scores of the following variables: percentage of overcrowded people; percentage of heads of household classified as “worker” or “employee”, “self-employed worker” or “family worker”; and percentage of people residing in low-quality housing. The level of social fragmentation was a modified version of the index developed by Congdon ^24^, calculated with the sum of z scores of the following variables: percentage of adults (over 17 years of age) single, divorced, legally separated or widowed; percentage of households made up of one person; percentage of households occupied by non-homeowners; and the percentage of the migrant population (people over 5 years of age, who 5 years before the census was carried out resided in another locality). During 2017-2021, annual variations were analyzed and the entire period was divided into two parts: pre-pandemic (2017-2019) and pandemic (2020-2021). Four regression models were applied. The first model estimated the relationship between the four categories of urbanization and suicide mortality during the entire period 2017-2021, controlling for sex, age, year of suicide occurrence, social fragmentation and poverty. The second model only differs from the first in its replacement of the year variable with a categorical variable distinguishing the pre-pandemic period from the pandemic period, due to the collinearity between both variables. The third model included the first in addition to an interaction between the urbanization variable and the year of suicide occurrence, to allow for the possibility of a differential evolution in suicide rates by urbanization category during the period 2017-2021. The fourth model included the second model in addition to an interaction between the urbanization variable and the period of suicide occurrence (pre-pandemic/pandemic). Additionally, these four models were calculated using SNIC data in two different ways. First, considering the suicides that occurred in 2019 and 2020 throughout the calendar year, in the same way as what was done with the MSAL data. Second, because the SNIC data contained the month of death, and that the first cases and deaths from COVID-19 began to be registered in the first days of March 2020, the pre-pandemic period extended during 2017 until February 2020, and the pandemic period during March 2020 until 2021. The data were analyzed using Stata version 13.1 (StataCorp, College Station, TX).

## Results

During the period 2017-2021, 4,980 and 4,897 suicides were reported according to the SNIC and the MSAL, respectively, considering cases with information on sex and age group. Regarding distribution by sex, both data sources were very similar (20% women and 80% men according to the SNIC, and 17%-83% according to the MSAL). Regarding the distribution by age group, we also found very similar values in populations aged 10-29 years (36% in SNIC versus 34% in MSAL), 30-59 years (42% in SNIC versus 44% in MSAL), and 60+ years (22% in SNIC versus 23% in MSAL). Figure 1 shows the geographical distribution of the population density quartiles, the social fragmentation index and the poverty index. The municipalities with lower population density (more rural) are located mainly in two large areas: central-east and the western part of the PBA (Figure 1, map A). On the other hand, the municipalities with the highest population density (more urbanized) are concentrated in the megacity Gran Buenos-La Plata (GBA). The social fragmentation index shows its highest values in municipalities of the central-eastern PBA (Figure 1, map B), while the poverty index mainly concentrates its highest values in municipalities of the GBA (Figure 1, map C).

**Figure 1.**
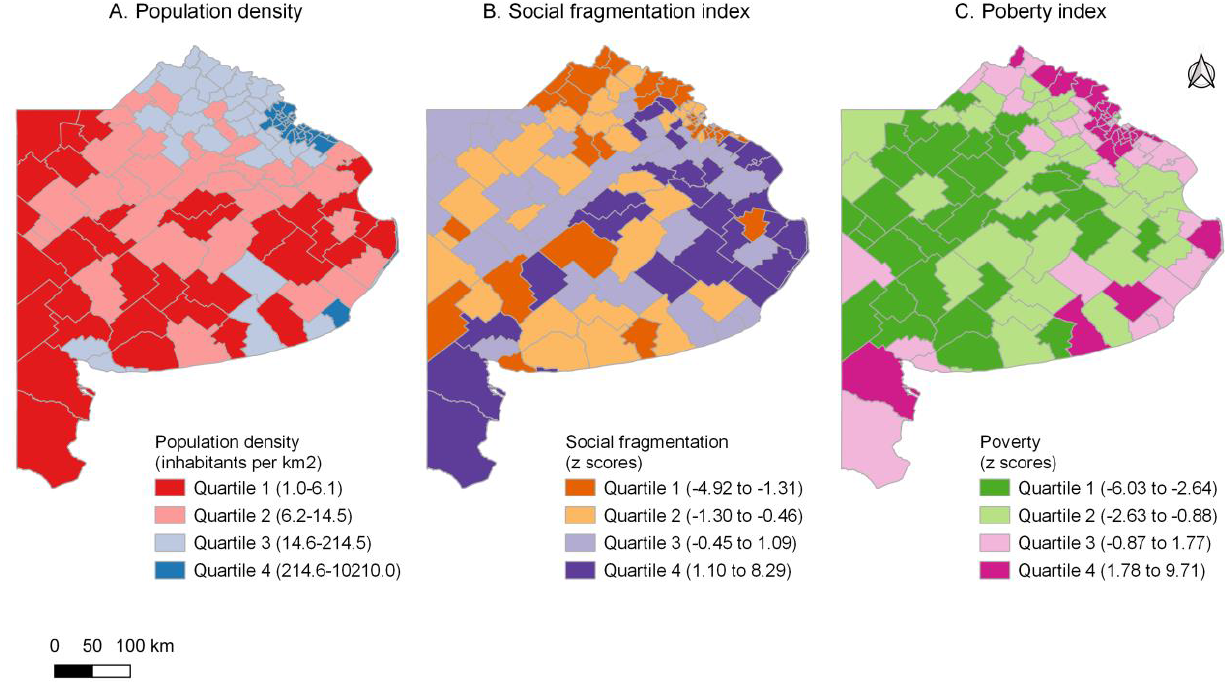
Geographic distribution of the quartiles of population density, social fragmentation and poverty in the Province of Buenos Aires, 2010.

Tables 1 and 2 and Figure 2 show the analyzes carried out with suicide data obtained from the MSAL. Considering models 1 and 2, male populations, aged 60+, residing in districts with high levels of social fragmentation, and with low levels of poverty presented a higher risk of suicide (Table 1). In the most urbanized municipalities, a lower risk of suicide was observed compared to the most rural municipalities. Considering model 1, there was a decrease in mortality in the period 2020-2021 compared to 2017 (Table 1). Model 2 showed a lower risk of suicide during the pandemic period compared to the pre-pandemic three-year period. When considering model 3, a downward trend in suicide rates can be observed towards the most urbanized quartiles, with relative risks – and confidence intervals – less than 1 in the most rural quartile, compared to the most urban quartile (Figure 2). Considering model 4, during the pre-pandemic period the most urbanized municipalities presented the lowest risk of suicide, compared to the most rural municipalities (Table 2). Relative urban-rural inequalities in suicide were similar between the pre-pandemic and pandemic periods.

**Table 1.**
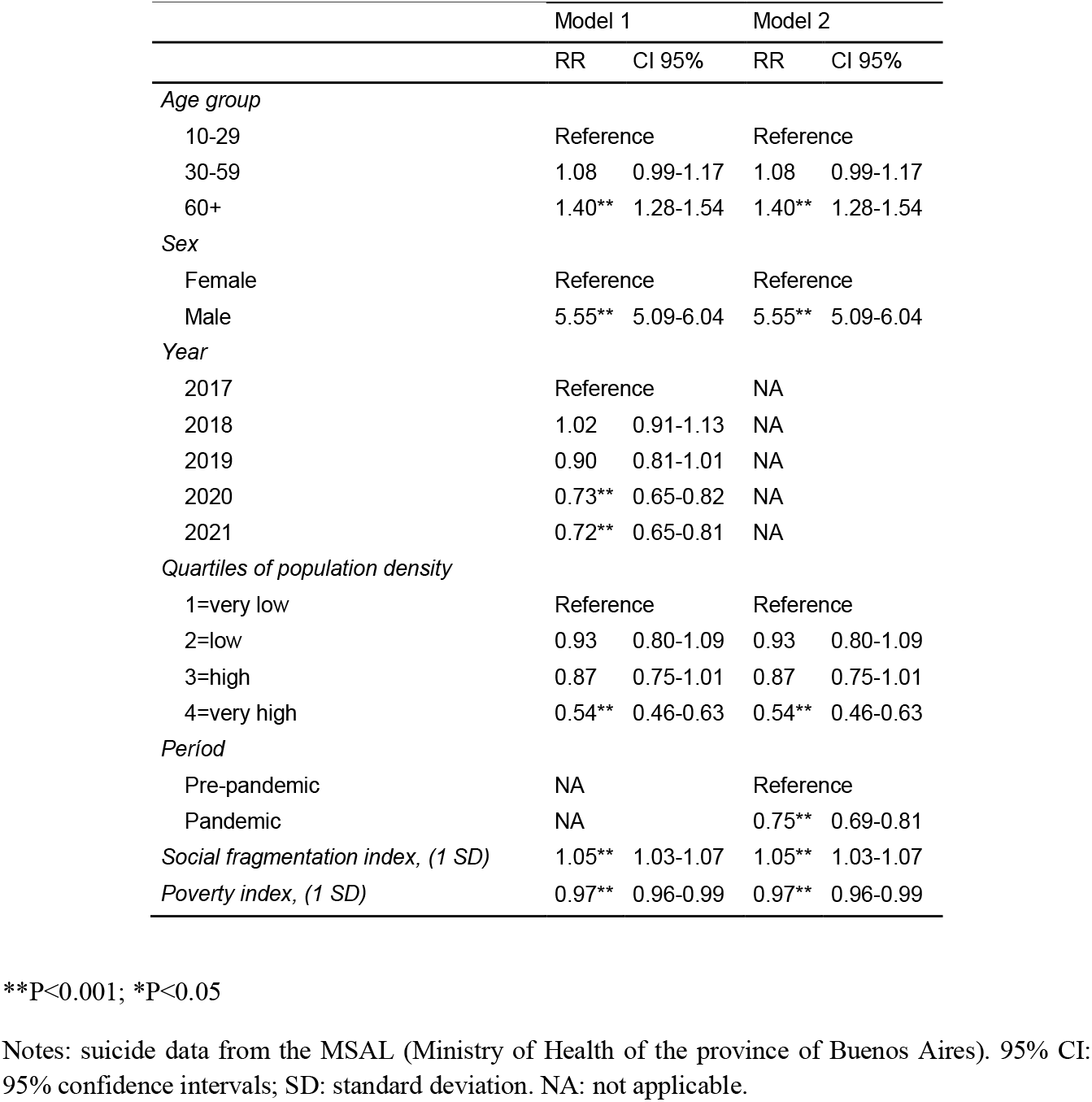
Relative risks (RR) of suicide associated with sex, age, year of occurrence (model 1), period (model 2), and sociodemographic characteristics of the municipalities. Province of Buenos Aires, 2017-2021. ^**^P<0.001; ^*^P<0.05

|  | Model 1 |  | Model 2 |  |
| --- | --- | --- | --- | --- |
|  | RR | CI 95% | RR | CI 95% |
| *Age group* |  |  |  |  |
| 10-29 | Reference |  | Reference |  |
| 30-59 | 1.08 | 0.99-1.17 | 1.08 | 0.99-1.17 |
| 60+ | 1.40** | 1.28-1.54 | 1.40** | 1.28-1.54 |
| *Sex* |  |  |  |  |
| Female | Reference |  | Reference |  |
| Male | 5.55** | 5.09-6.04 | 5.55** | 5.09-6.04 |
| *Year* |  |  |  |  |
| 2017 | Reference |  | NA |  |
| 2018 | 1.02 | 0.91-1.13 | NA |  |
| 2019 | 0.90 | 0.81-1.01 | NA |  |
| 2020 | 0.73** | 0.65-0.82 | NA |  |
| 2021 | 0.72** | 0.65-0.81 | NA |  |
| *Quartiles of population density* |  |  |  |  |
| 1=very low | Reference |  | Reference |  |
| 2=low | 0.93 | 0.80-1.09 | 0.93 | 0.80-1.09 |
| 3=high | 0.87 | 0.75-1.01 | 0.87 | 0.75-1.01 |
| 4=very high | 0.54** | 0.46-0.63 | 0.54** | 0.46-0.63 |
| *Period* |  |  |  |  |
| Pre-pandemic | NA |  | Reference |  |
| Pandemic | NA |  | 0.75** | 0.69-0.81 |
| *Social fragmentation index, (1 SD)* | 1.05** | 1.03-1.07 | 1.05** | 1.03-1.07 |
| *Poverty index, (1 SD)* | 0.97** | 0.96-0.99 | 0.97** | 0.96-0.99 |

\*\*P<0.001; \*P<0.05
Notes: suicide data from the MSAL (Ministry of Health of the province of Buenos Aires). 95% CI: 95% confidence intervals; SD: standard deviation. NA: not applicable.

**Table 2.** Rates (per 100,000 inhabitants) and relative risks of suicide associated with quartiles of population density and during the pre-pandemic and pandemic periods. Province of Buenos Aires, 2017-2021. ^**^P<0.001; ^*^P<0.05

| Period | Population density | Suicide rate | Relative risk |
| --- | --- | --- | --- |
| Pre-pandemic | 1=very low | 13.76 (11.62 - 15.90) | Reference |
|  | 2=low | 12.47 (10.94 - 14.01) | 0.91 (0.75 - 1.10) |
|  | 3=high | 12.36 (11.22 - 13.50) | 0.90 (0.75 - 1.08) |
|  | 4=very high | 7.86 (7.22 - 8.51) | 0.57 (0.48 - 0.69)** |
| Pandemic | 1=very low | 11.34 (9.02 - 13.67) | Reference |
|  | 2=low | 11.11 (9.38 - 12.83) | 0.98 (0.76 - 1.26) |
|  | 3=high | 9.35 (8.20 - 10.49) | 0.82 (0.65 - 1.05) |
|  | 4=very high | 5.48 (4.94 - 6.02) | 0.48 (0.38 - 0.61)** |

\*\*P<0.001; \*P<0.05
Notes: suicide data from the MSAL (Ministry of Health of the province of Buenos Aires). In parentheses, 95% confidence intervals. Rates and relative risks estimated considering model 4.

**Figure 2.**
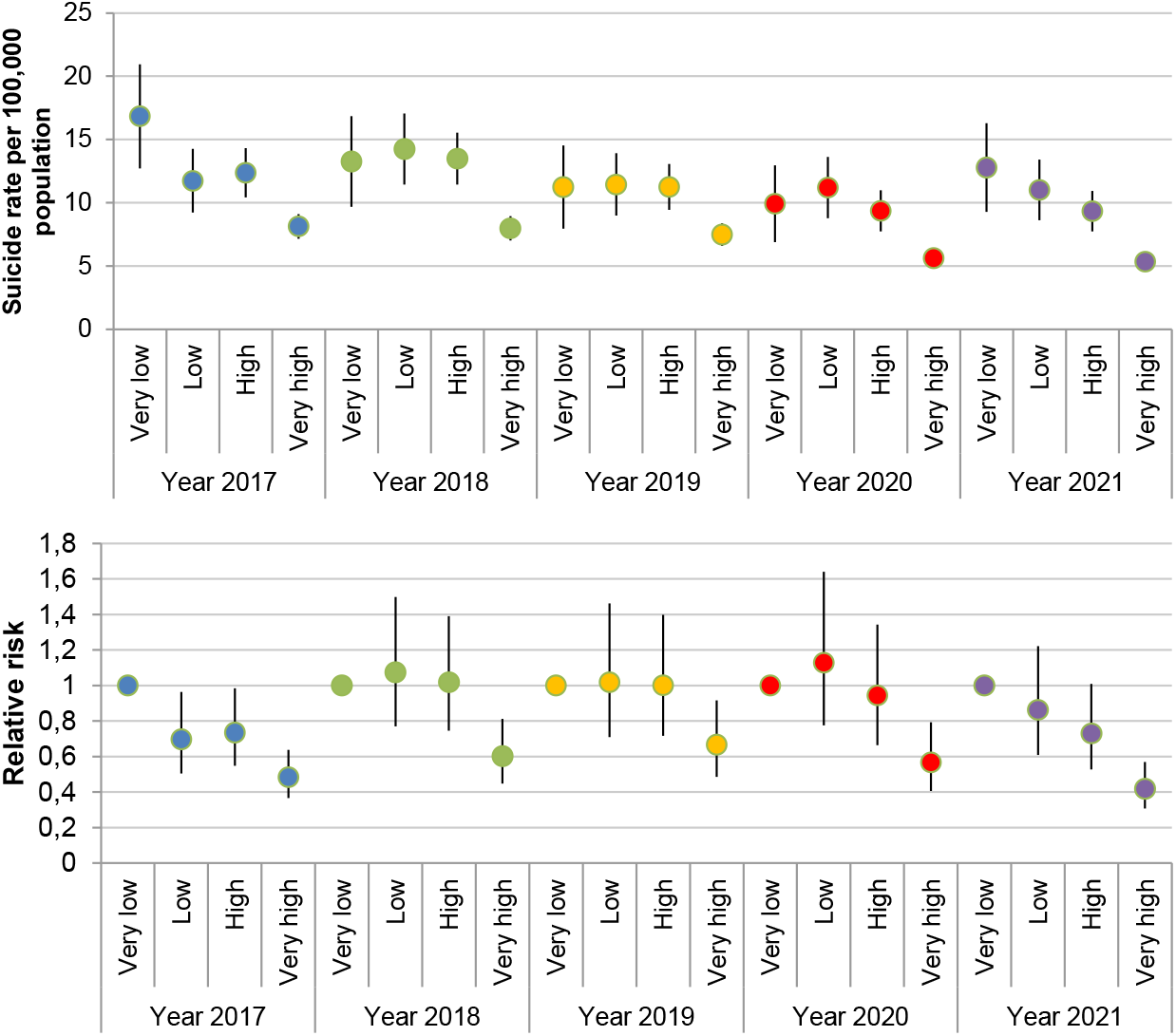
Rates (per 100,000 inhabitants, upper panel) and relative risks (lower panel) of suicide according to each year, quartile of population density (“Very low” = lowest density quartile; “Very high” = quartile of the highest density). Province of Buenos Aires, 2017-2021. Notes: rates and relative risks estimated considering model 3. In the case of relative risks, the reference is the quartile of lowest population density of each year, so model 3 was calculated five times changing the reference year in its interaction with population density quartiles. Vertical lines indicate 95% confidence intervals. Suicide data from the MSAL (Ministry of Health of the province of Buenos Aires).

Overall, analyzes with SNIC data showed similar results compared to MSAL data, although some differences were observed. Considering model 1, the risk of suicide only decreased in 2020 compared to 2017 (Table 1A, Supplementary Material). With respect to model 3, in 2020 no relative inequalities in the risk of suicide were observed between urbanization quartiles, due to a much lower suicide rate in the quartile with the lowest population density, compared to the rate calculated based on data from the MSAL (Figure 1A, Supplementary Material). Similarly, considering model 4, during the pandemic period there was no difference in the relative risk of suicide between the most rural and the most urbanized municipalities (Table 2A, Supplementary material). This was due to a marked decrease in the suicide rate in the most rural municipalities, indicating a higher level of under-registration of suicides when compared to the rate calculated with data from the MSAL in these municipalities.

Finally, the results of the regression models using SNIC data, but considering all suicides reported within each of the years 2019 and 2020, were similar to those obtained considering the pre-pandemic period until February 2020 and the pandemic period starting in March 2020. Regarding model 1, by not including suicides reported during January and February 2020, the year 2019 had a lower risk of suicide compared to 2017 (Table 1A, Supplementary material). Except for this difference, both models using SNIC data are very similar. In model 2, the pandemic period was associated with a lower risk of suicide considering it both from January 2020 and from March 2020 (Table 1A, Supplementary material). Similar results were also found in models 3 (Figure 2A, Supplementary Material) and 4 (Table 2A, Supplementary Material).

## Discussion

This study used the most recent suicide data in the PBA, finding a higher risk of suicide in rural areas, compared to more urbanized areas, after controlling for indices of social fragmentation and poverty, in line with a previous study in Argentina ^5^. Using two data sources, or considering two different beginnings of the pandemic period (with data from the SNIC), our results showed a lower risk of suicide during this period compared to the pre-pandemic period. Finally, urban-rural suicide gaps did not increase with the onset of the pandemic.

On the other hand, we found some differences in the temporal and urban-rural variations of suicide using the two data sources, which could be due to two factors. First, vital statistics - MSAL-seem to underestimate suicides in the most urbanized districts. Second, there seemed to be a significant under-reporting of suicides in the SNIC data in the most rural districts during 2020. Despite these differences, both data sources showed very similar types and levels of association with both sex and age as well as with the indices of social fragmentation and poverty.

If we consider the main differences between both sources of data due to under-registration of suicides, the general stability in the relative inequalities between the most rural and the most urbanized municipalities seems to indicate that the pandemic did not intensify the factors that increase the risk of suicide in rural areas, as was believed at the beginning of the COVID-19 pandemic ^15^.

During the first wave of COVID-19 cases, in 2020, mobility restrictions between cities in the PBA may have reduced the uncertainty of contagion in rural populations, by hindering the hierarchical spread of the virus from Greater Buenos Aires. At the same time, because economic activities linked to agricultural production and distribution, predominant in municipalities with low population density, were declared essential during the COVID-19 pandemic, it is likely that the impact of social isolation measures has been lower in rural areas compared to the largest cities in the PBA. Another factor is the greater availability of green spaces in municipalities with lower population density and the very low proportion of households residing in apartments –generally without availability of green space within the housing property–, compared to municipalities with higher population density. These factors could have generated less feeling of confinement and isolation in rural areas. Furthermore, together with the geographical isolation, they could contribute to generating a feeling of security in the populations of the most rural municipalities ^25^.

Another effect of the beginning of the COVID-19 pandemic, and the subsequent establishment of restrictions on population mobility, was the retention of the young population in towns and small cities that annually travel to larger cities to attend university. It is possible that this brief retention of the young population in the most rural districts during 2020 has intensified family ties in some households, acting as a protective factor against suicide. According to Baudelot and Establet ^26^, and following Durkheim ^13^, the risk of suicide decreases when the level of social integration increases, understood as the intensity and quantity of ties that an individual establishes with his or her family environment.

This study has several limitations. First, as mentioned above, both data sources would present limitations associated with the under-reporting of suicides. With respect to the SNIC data, it is possible that this under-registration is the cause of the decrease in suicides in the most rural districts during 2020. Within the municipalities of the quartile with the lowest population density, when analyzing the differences between the average number of suicides registered during 2017-2019 and suicides reported in 2020 (data not shown in Results), the greatest differences occur mainly in municipalities in the southwest of the PBA (Coronel Dorrego, Patagones, Villarino), close to the city of Bahía Blanca, with the highest population size in that area of the PBA. If there was under-registration in these municipalities with low population density, an additional hypothesis could be related to movements of police officers from these municipalities, during 2020, to the city of Bahía Blanca. However, it is only one possible explanation for which we do not have empirical support.

Regarding the MSAL data, it is possible an under-reporting of suicides coded as deaths due to injuries of undetermined intent. However, evidence in Argentina shows that a majority of these deaths would actually correspond to homicides ^27,28^. Second, we used census projections based on the variation in the population, by sex and age groups, between the 2001 and 2010 censuses. Therefore, these projections may present discrepancies with the data from the census carried out in 2022. Unfortunately, this information is not yet available, which prevented us from estimating rates and relative risks of suicide in a more precise way.

In conclusion, our results do not support the hypothesis of a worsening of the factors that, in rural areas, increase the risk of suicide during the COVID-19 pandemic period. However, a high risk of suicide persisted in these areas, making it imperative to implement public policies aimed at reversing rural depopulation and increasing access to mental health services.

## Data Availability

All data produced are available online at:
https://catalogo.datos.gba.gob.ar/dataset/defunciones-generales
https://www.argentina.gob.ar/seguridad/estadisticascriminales

## Conflict of Interest

All authors declare no conflict of interest.

Ethics committee approval was not necessary because we used secondary data that was freely accessible to any Argentine citizen.

