## Supplementary material for "COVID-19 pandemic and changes in urban-rural inequalities of suicide in the Province of Buenos Aires-Argentina, 2017-2021"

**Table 1A**. Relative risks (RR) of suicide associated with sex, age, year of occurrence (model 1), period (model 2), and sociodemographic characteristics of the municipalities. Province of Buenos Aires, 2017-2021.

| | SNIC[‡] | | SNIC-2[§] | |
|---|---|---|---|---|
| | RR | CI 95% | RR | CI 95% |
| Model 1 | | | | |
| *Age group* | | | | |
| 10-29 | Reference | | Reference | |
| 30-59 | 0.98 | 0.90-1.06 | 0.98 | 0.90-1.06 |
| 60+ | 1.28** | 1.17-1.40 | 1.27** | 1.17-1.39 |
| *Sex* | | | | |
| Female | Reference | | Reference | |
| Male | 4.49** | 4.15-4.86 | 4.48** | 4.14-4.85 |
| *Year* | | | | |
| 2017 | Reference | | Reference | |
| 2018 | 1.32** | 1.19-1.46 | 1.32** | 1.19-1.46 |
| 2019 | 1.00 | 0.90-1.11 | 0.87* | 0.78-0.97 |
| 2020 | 0.63** | 0.56-0.70 | 0.75** | 0.67-0.84 |
| 2021 | 0.96 | 0.86-1.07 | 0.96 | 0.86-1.07 |
| *Quartiles of population density* | | | | |
| 1=very low | Reference | | Reference | |
| 2=low | 1.08 | 0.91-1.28 | 1.08 | 0.91-1.27 |
| 3=high | 0.97 | 0.83-1.13 | 0.97 | 0.83-1.13 |
| 4=very high | 0.73** | 0.63-0.86 | 0.73** | 0.62-0.86 |
| *Social fragmentation index, (1 SD[⁋])* | 1.06** | 1.04-1.08 | 1.06** | 1.04-1.08 |
| *Poverty index, (1 SD[⁋])* | 0.97** | 0.96-0.99 | 0.97** | 0.96-0.99 |
| Model 2 | | | | |
| *Age group* | | | | |
| 10-29 | Reference | | Reference | |
| 30-59 | 0.97 | 0.90-1.06 | 0.97 | 0.90-1.06 |
| 60+ | 1.27** | 1.16-1.40 | 1.27** | 1.16-1.40 |
| *Sex* | | | | |
| Female | Reference | | Reference | |
| Male | 4.50** | 4.15-4.88 | 4.51** | 4.15-4.89 |
| *Quartiles of population density* | | | | |
| 1= very low | Reference | | Reference | |
| 2=low | 1.08 | 0.91-1.28 | 1.08 | 0.91-1.28 |
| 3=high | 0.97 | 0.83-1.14 | 0.97 | 0.83-1.14 |
| 4=very high | 0.74** | 0.63-0.87 | 0.74** | 0.63-0.87 |
| *Períod* | | | | |
| Pre-pandemic | Reference | | Reference | |
| Pandemic | 0.72** | 0.67-0.77 | 0.80** | 0.75-0.86 |

| | | | | |
|---|---|---|---|---|
| *Social fragmentation index, (1 SD[⁋])* | 1.06** | 1.04-1.08 | 1.06** | 1.04-1.08 |
| *Poverty index, (1 SD[⁋])* | 0.97** | 0.96-0.99 | 0.97** | 0.96-0.99 |

**P<0.001; *P<0.05

Note: suicide data from the SNIC (National Criminal Information System of the Ministry of National Security). 95% CI: 95% confidence intervals.

[‡] Suicides in 2019 also include those that occurred during January and February 2020, while in 2020 the suicides that occurred between March (first reported cases of COVID-19 in Argentina) and December were considered.

[§] In both 2019 and 2020, suicides reported between January and December were considered.

[⁋] SD: standard deviation.

**Table 2A**. Rates (per 100,000 inhabitants) and relative risks* of suicide associated with quartiles of population density and during the pre-pandemic and pandemic periods. Province of Buenos Aires, 2017-2021.

| | | SNIC data ‡ | | SNIC-2 data § | |
|---|---|---|---|---|---|
| Period | Population density | Suicide rate | Relative risk | Suicide rate | Relative risk |
| Pre-pandemic | 1=very low | 12.15 (10.18 - 14.13) | Reference | 11.68 (9.75 - 13.62) | Reference |
| | 2=low | 11.74 (10.28 - 13.20) | 0.97 (0.79 - 1.18) | 11.37 (9.94 - 12.81) | 0.97 (0.79 - 1.20) |
| | 3=high | 10.86 (9.84 - 11.88) | 0.89 (0.74 - 1.08) | 10.31 (9.33 - 11.30) | 0.88 (0.73 - 1.07) |
| | 4=very high | 8.53 (7.85 - 9.20) | 0.70 (0.58 - 0.85)** | 8.23 (7.57 - 8.89) | 0.70 (0.58 - 0.85)** |
| Pandemic | 1=very low | 7.13 (5.32 - 8.94) | Reference | 7.82 (5.92 - 9.71) | Reference |
| | 2=low | 9.73 (8.15 - 11.30) | 1.36 (1.01 - 1.84)* | 10.25 (8.62 - 11.87) | 1.31 (0.98 - 1.75) |
| | 3=high | 8.26 (7.22 - 9.30) | 1.16 (0.87 - 1.54) | 9.06 (7.96 - 10.16) | 1.16 (0.88 - 1.52) |
| | 4=very high | 5.89 (5.33 - 6.45) | 0.83 (0.63 - 1.09) | 6.33 (5.74 - 6.92) | 0.81 (0.62 - 1.05) |

**P<0.001; *P<0.05

Notes: rates and relative risks estimated considering model 4. Suicide data from the SNIC (National Criminal Information System of the Ministry of National Security). In parentheses, 95% confidence intervals.

‡ Suicides in 2019 also include those that occurred during January and February 2020, while in 2020 suicides that occurred between March (first reported cases of COVID-19 in Argentina) and December were considered.

§ In both 2019 and 2020, suicides reported between January and December were considered.

**Figure 1A.** Rates (per 100,000 inhabitants, upper panel) and relative risks (lower panel) of suicide according to each year, quartile of population density (“Very low” = lowest density quartile; “Very high” = highest density quartile ), and considering the year 2020 starting in March (SNIC) and starting in January (SNIC-2). Province of Buenos Aires, 2017-2021.

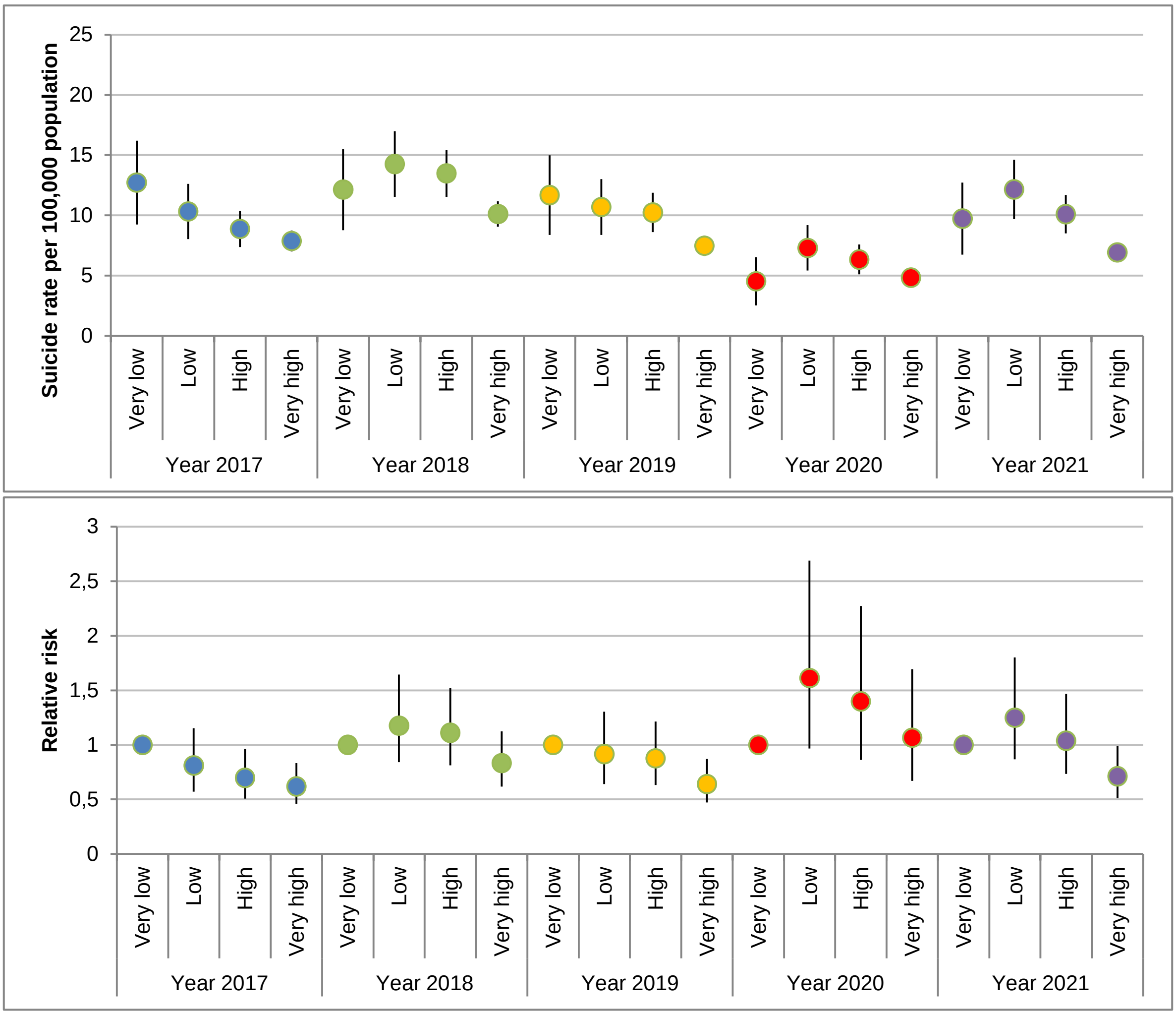


Notes: rates and relative risks estimated considering model 3. In the case of relative risks, the reference is the quartile of lowest population density of each year, so model 3 was calculated five times changing the reference year in its interaction with population density quartiles. Vertical lines indicate 95% confidence intervals. Suicide data from the SNIC.

**Figure 2A**. Rates (per 100,000 inhabitants, upper panel) and relative risks (lower panel) of suicide according to each year, quartile of population density (“1”=lowest density quartile; “4”=highest density quartile), and considering the year 2020 starting in March (SNIC) and starting in January (SNIC-2). Province of Buenos Aires, 2017-2021.

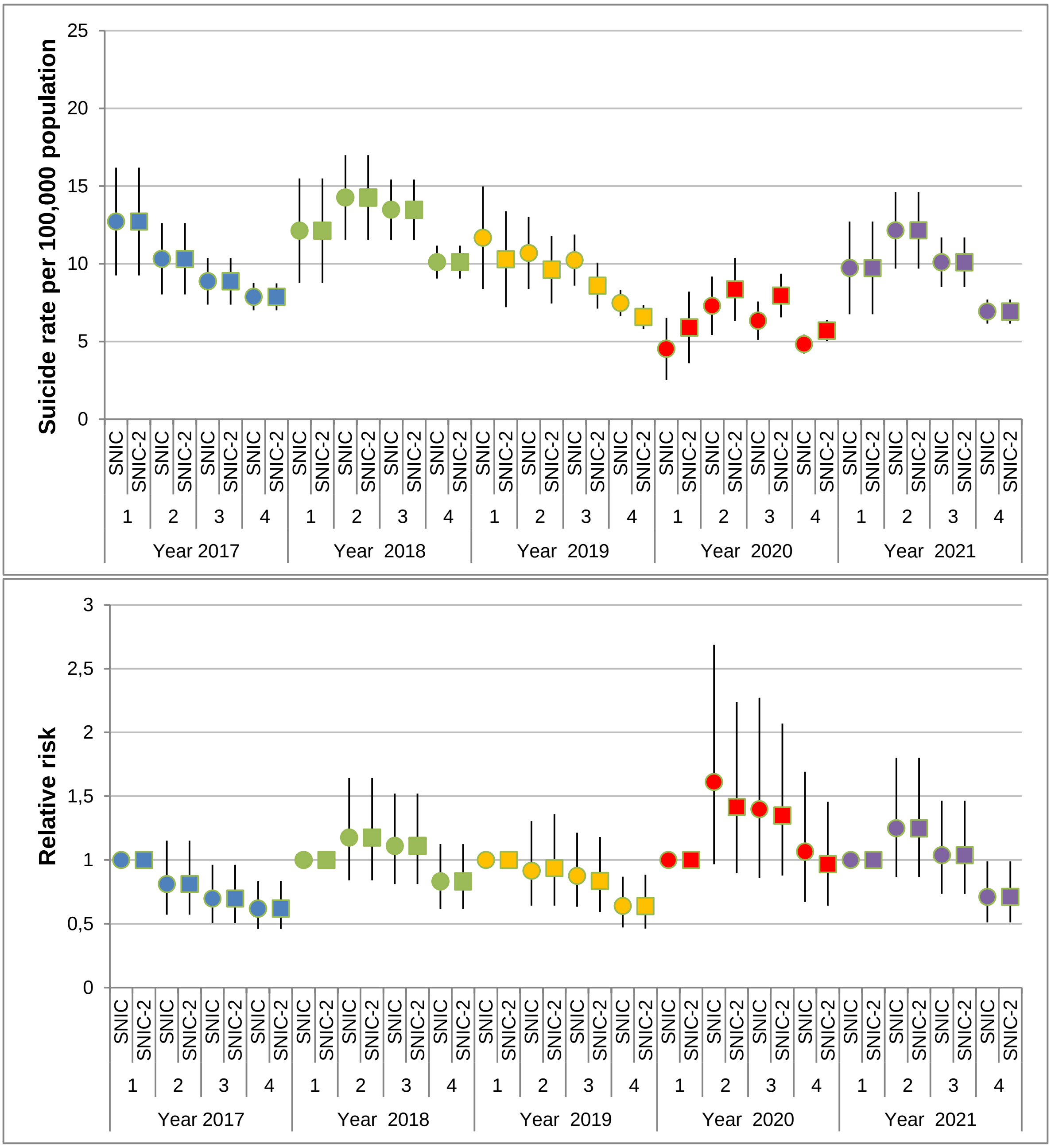


Notes: rates and relative risks estimated considering model 3. In the case of relative risks, the reference is the quartile of lowest population density of each year, so model 3 was calculated five times changing the reference year in its interaction with population density quartiles. Vertical lines indicate 95% confidence intervals.
